# Accuracy of Artificial Intelligence-Based Models versus Traditional Scoring Systems (APACHE, SOFA, SAPS) for Predicting Mortality in ICU Patients: A Systematic Review and Meta-Analysis

**DOI:** 10.64898/2026.01.14.26344000

**Authors:** Vimukta Pradhan, Himanshu Shekhar, Punam Kumari Munda, Ashutosh Kumar Tiwari, Sneha Jha, Pratibha Rai

## Abstract

**Introduction:** Reliable estimation of mortality among critically ill patients is crucial for guiding clinical decisions and optimizing ICU performance. Traditional scoring systems such as APACHE, SOFA, and SAPS are commonly applied, though their predictive capacity is constrained by their reliance on static structures and linear modeling assumptions. Artificial intelligence-based models provide flexible, data-oriented prediction strategies, yet their comparative accuracy remains unclear. This study systematically reviewed and meta-analyzed the performance of Artificial intelligence-based models versus conventional ICU scores for predicting in-hospital mortality in adults admitted to ICU.

**Materials and Methods:** Literature searches were performed in PubMed, Embase, Web of Science, Scopus and the Cochrane Library from January 2015 to August 2025 for studies comparing AI models with traditional scoring systems. Studies were included if they provided diagnostic performance indicators including AUC, sensitivity, or specificity. Risk of bias was assessed using PROBAST, and pooled statistical estimates were derived through bivariate random-effects modeling with Fisher’ s *Z-* transformation. Subgroup analyses examined AI modality, ICU type, and geographic region.

**Results:** Eleven studies involving over one million ICU admissions met inclusion criteria. Two studies (Huang 2023; Lim 2024) provided complete 2 by 2 data for meta-analysis. Pooled sensitivity and specificity for AI models were 0.875 (95% CI: 0.840 - 0.904) and 0.857 (95% CI: 0.845 - 0.868), respectively. AI models achieved higher AUCs (0.82 - 0.90) than APACHE II (0.70 - 0.78), SOFA (0.68 - 0.75), and SAPS II (0.70 - 0.79). Deep learning and ensemble methods performed best across ICU settings and regions.

**Conclusion:** AI-based models outperform conventional scoring systems in predicting ICU mortality. Their integration into critical care could enhance early risk stratification and precision prognostication.

**Highlights:** This meta-analysis highlights that artificial intelligence–based predictive models demonstrated superior predictive performance than conventional ICU scoring systems (APACHE, SOFA, and SAPS) in predicting in-hospital mortality, with higher pooled sensitivity, specificity, and overall discriminative accuracy, particularly for deep learning and ensemble approaches across diverse ICU settings.

## INTRODUCTION

Mortality prediction remains a core challenge in critical care units, where timely and accurate prognostication directly influences the clinical outcomes, resource utilization, and overall quality of care in intensive care medicine. Accurate prediction of mortality among critically ill patients continues to be a cornerstone of intensive care medicine. Early identification of individuals with critical illness who are at high risk of mortality enables clinicians to optimize resource allocation more efficiently, guide treatment strategies, and maintain quality benchmarking across intensive care units (ICUs).^1,2^ Over the past few decades, conventional scoring systems such as the Acute Physiology and Chronic Health Evaluation (APACHE), Simplified Acute Physiology Score (SAPS), and Sequential Organ Failure Assessment (SOFA) have been widely used to predict outcomes among patients admitted across various ICUs worldwide.^3,4^ These models typically depend physiological, biochemical, and demographic variables collected early after ICU admission, usually within the first 24 hours post admission to ICU to provide the mortality risk estimate. Despite their widespread adoption, the predictive performance of these scoring systems varies across populations and healthcare settings, frequently necessitating recalibration to retain validity.^5,6^

As ICU populations evolve and real-time data become increasingly available, traditional scoring systems—largely based on classical regression techniques and predefined clinical variables—are often considered too static to capture the dynamic complexity of contemporary critical care. While this approach offers transparency, it assumes additive and independent effects among predictors, potentially overlooking complex nonlinear relationships that influence mortality.^7,8^ Moreover, these models are inherently static, limiting their ability to incorporate continuous patient monitoring data or adapt to temporal changes in physiological status and biochemical parametres.^9^ Consequently, their discrimination and calibration often deteriorate over time, particularly as therapeutic practices and diverse patient characteristics evolve.^10^ Previous studies have indicated that APACHE II and SAPS II demonstrate moderate discrimination, with areas under the curve (AUCs) generally ranging from 0.70 to 0.80; however, performance often declines when these models are applied to external or specialized ICU cohorts.^9–11^ These limitations have driven the pursuit of more adaptive and sophisticated approaches to mortality prediction in critical care management.

The growing application of artificial intelligence (AI) and machine learning (ML) has ushered in a new generation of predictive modelling in critical care.^12^ By leveraging large, high-dimensional datasets from electronic health records (EHRs), laboratory investigations, and continuous patient-monitoring systems, AI-based algorithms are able to identify complex non-linear relationships that frequently elude traditional statistical approaches.^13^ Techniques such as random forests, gradient boosting machines, and deep learning models—including convolutional neural networks (CNNs) and long short-term memory (LSTM) networks—have shown strong potential to integrate diverse clinical information effectively.^14^ Studies utilizing large databases such as MIMIC-III, the eICU Collaborative Research Database, and institutional EHR repositories frequently reported that AI-based models demonstrate superior predictive accuracy in comparison with conventional scoring systems.^15^ The inherent adaptability of AI allows continuous recalibration, thereby enhancing model generalizability and sustaining predictive reliability over time.

Despite these advances, the existing body of evidence remains fragmented and, in some instances, contradictory. Studies vary widely in their design, dataset size, feature selection, and modelling approach, resulting in inconsistent estimates of predictive performance.^16^ While several studies have underscored clear performance gains of AI-based models over conventional scores, many others have shown only marginal improvements or insignificant difference when validated externally.^17^ The use of diverse performance metrics—ranging from accuracy and AUC to sensitivity and specificity—further complicates any direct comparison across studies. Moreover, potential sources of bias related to patient selection, overfitting, and incomplete reporting of model development continue to pose challenges.^18^ Most of the earlier systematic reviews appeared before deep learning and ensemble methods became common, and they often overlooked a structured assessment of methodological quality using frameworks such as PROBAST, which are now regarded as essential for prediction-model evaluations.^19^

Although research on artificial intelligence (AI) in critical care has expanded rapidly in recent years, comparisons with conventional ICU scoring systems remain limited, with substantial variation in study design, sample size, and validation strategy leading to inconsistent estimates of predictive accuracy. Moreover, previous reviews have often failed to incorporate recent advances in deep learning and ensemble modelling or to apply standardized tools such as PROBAST for methodological quality assessment. These limitations make it difficult for clinicians to interpret the true value of AI models in routine ICU practice. Therefore, this systematic review and meta-analysis was carried out to provide an updated and methodologically robust synthesis of evidence comparing AI-based predictive models with traditional ICU scoring systems (APACHE, SOFA, and SAPS) for predicting in-hospital mortality in adult patients, thereby addressing existing evidence gaps and supporting more informed integration of AI tools into critical care prognostication.

## MATERIALS AND METHODS

### Study Design and Registration

This systematic review and meta-analysis was conducted in accordance with the Preferred Reporting Items for Systematic Reviews and Meta-Analyses (PRISMA 2020).^20^

The review protocol was registered prospectively with the International Prospective Register of Systematic Reviews (PROSPERO) under registration number CRD420251168073.

### Objectives

The primary objective of this systematic review and meta-analysis was to provide an updated and methodologically robust comparison of AI-based predictive models with conventional ICU scoring systems—namely APACHE (II/IV), SOFA, and SAPS (II)—for predicting in-hospital mortality among adult critically ill patients. Secondary objectives were to assess the diagnostic performance of these models by analyzing metrics such as sensitivity, specificity, and area under the curve (AUC); to examine study quality and potential bias through the PROBAST assessment tool; and to perform subgroup analyses based on AI approach, ICU setting, and geographical location to investigate heterogeneity and determinants of model effectiveness.

### Eligibility Criteria

Eligibility criteria were defined according to the PICOS framework, consistent with the study registration in PROSPERO.

Population (P): Adult patients (≥18 years) admitted to medical, surgical, or mixed intensive care units.

Intervention (I): AI-based or ML–based models developed or externally validated for predicting in-hospital or ICU mortality.

Comparator (C): Traditional ICU scoring systems, including APACHE II/IV, SOFA, or SAPS II, were assessed within the same study cohort.

Outcomes (O): Primary outcome—Diagnostic accuracy for mortality prediction, expressed as sensitivity, specificity, and area under the receiver operating characteristic curve (AUC). Secondary outcomes—Subgroup analyses by AI type, ICU setting, and geographic region; evaluation of study quality and bias using the PROBAST tool.

Study Design (S): Retrospective or prospective cohort studies directly comparing AI-based models with conventional scoring systems and reporting diagnostic performance metrics were eligible.

### Inclusion Criteria

Studies were included if they:

- Involved adult ICU populations (≥18 years).
- Evaluated or validated AI/ML models for predicting in-hospital or ICU mortality.
- Included a comparator scoring system (APACHE II/IV, SOFA, or SAPS II).
- Reported diagnostic accuracy measures (AUC, sensitivity, specificity) or data permitting their calculation.
- Were peer-reviewed, published

### Exclusion Criteria

Studies were excluded if they:

- Included pediatric populations (<18 years) or animal models.
- Used simulated or non-clinical datasets.
- Did not include mortality as an outcome.
- Lacked a comparator traditional scoring system.
- Failed to report diagnostic accuracy measures (AUC, sensitivity, specificity, or data to construct 2×2 tables).
- Were reviews, commentaries, protocols, abstracts, or non–peer-reviewed publications.

### Search Strategy

A comprehensive literature search was conducted across PubMed, Embase, Scopus, Web of Science, and the Cochrane Library for studies published between January 2015 and August 2025.

Both controlled vocabulary (MeSH terms) and free-text keywords were used, combined with Boolean operators to identify relevant studies on ICU mortality, AI, and conventional scoring systems.

The detailed PubMed search strategy was (“Intensive Care Units” [MeSH] OR “critical illness” OR “ICU”) AND (“mortality” OR “in-hospital mortality” OR “death”) AND (“artificial intelligence” OR “machine learning” OR “deep learning” OR “neural network” OR “predictive model”) AND (“APACHE” OR “SOFA” OR “SAPS” OR “scoring system”).

Equivalent search strategies, adapted to the indexing terms and syntax of each database, were applied to Embase, Scopus, Web of Science, and the Cochrane Library.

The reference sections of all included articles and pertinent reviews were manually examined to locate any additional studies meeting the inclusion criteria. Only peer-reviewed studies published in English Language were included.

### Data Extraction

Two independent reviewers extracted data on study characteristics, cohort details, AI model type, comparator scoring system, validation method, sample size, and performance metrics (AUC, sensitivity, specificity).

Any discrepancies between reviewers were settled through discussion, and when consensus could not be reached, a third reviewer provided arbitration. Where possible, data on true positives (TP), false positives (FP), false negatives (FN), and true negatives (TN) were extracted or computed to derive pooled diagnostic performance measures. Studies reporting only AUC values were included in the quantitative synthesis of discrimination performance.

### Quality Assessment

The risk of bias, methodological quality, and applicability of the included studies were assessed using the Prediction Model Risk of Bias Assessment Tool (PROBAST).

This tool assesses four domains: participants, predictors, outcomes, and analysis. Each study was independently evaluated for adequacy of cohort selection, timing and consistency of predictor measurement, clarity of outcome definition, and appropriateness of statistical methods.

PROBAST was selected because it is specifically designed for evaluating prediction model development and validation studies, including those employing machine learning and deep learning architectures.

### Statistical Analysis

Statistical analyses were performed using Review Manager (RevMan) version 5.4 and R software (metaDTA package). For studies reporting complete 2×2 contingency data, pooled sensitivity, specificity, and summary receiver operating characteristic (SROC) curves were estimated using a bivariate random-effects model (Reitsma et al.) with 95% confidence intervals (CIs). For studies reporting only area under the curve (AUC) values, pooled AUC estimates were derived using a random-effects meta-analysis with Fisher’s *Z*-transformation. All analyses were conducted using MetaDisc and RevMan version 5.4 to account for between-study variability. When available, true-positive, false-positive, false-negative, and true-negative values were used to compute diagnostic accuracy parameters. Studies with incomplete contingency data were retained for qualitative synthesis but excluded from quantitative pooling.

Between-study heterogeneity was assessed using τ², Cochran’s Q, and I² statistics, with I² values > 50% indicating substantial heterogeneity. Potential sources of heterogeneity were examined through predefined subgroup analyses based on AI modality (e.g., deep learning, ensemble, or traditional ML models), ICU type (medical, surgical, or mixed), and geographic region. Sensitivity analyses were performed by sequentially excluding studies with unclear or high risk of bias on the PROBAST assessment to evaluate the robustness of pooled estimates. Publication bias was not formally assessed because of the limited number of eligible studies. Assessment of reporting bias was planned using visual inspection of funnel plot asymmetry and Deeks’ regression test, as recommended for diagnostic accuracy meta-analyses. However, these analyses were not undertaken because fewer than ten studies contributed to each quantitative synthesis, making such assessments statistically unreliable. Risk of bias within individual studies was evaluated separately using the PROBAST tool, as described in the Quality Assessment subsection.

### Data Sources and Final Synthesis

The final synthesis incorporated 11 studies that met the eligibility criteria, representing a combined cohort of over one million ICU admissions. Two studies (Huang et al., 2023; Lim et al., 2024) provided complete diagnostic contingency data suitable for quantitative pooling, whereas the remaining studies contributed AUC-based performance metrics for qualitative synthesis.

Results from all included studies were summarized in structured tables describing study characteristics, AI model type, comparator scoring systems, and reported diagnostic performance. Quantitative synthesis results were presented using forest plots for pooled sensitivity and specificity, along with a summary receiver operating characteristic (SROC) curve illustrating overall discriminative performance. These tabular and graphical representations were developed to provide a clear and consistent comparison of AI-based predictive models with conventional ICU scoring systems across diverse settings.

## RESULTS

The initial search across five electronic databases and manual reference screening identified 196 records. After removal of 68 duplicates, 128 titles and abstracts were screened, yielding 96 full-text articles for assessment. Of these, 83 were successfully retrieved and reviewed in detail, and 72 were excluded for reasons including incomplete accuracy data, use of non-AI models, inclusion of pediatric populations, or irrelevant outcomes. Ultimately, 11 studies met the inclusion criteria for qualitative synthesis, while two studies (Huang 2023; Lim 2024) provided complete diagnostic contingency data suitable for inclusion in the quantitative diagnostic test accuracy meta-analysis.

The PRISMA 2020 flow diagram (Figure 1) summarizes the study selection process for this systematic review and meta-analysis. The included studies, published between January 2016 and August 2025, collectively analyzed data from more than one million ICU admissions across diverse geographic regions, including North America, Europe, and Asia. Sample sizes in individual studies ranged from approximately 5,200 to over 480,000 patients, with mean participant ages between 55 and 68 years. These studies encompassed a variety of ICU populations and clinical contexts, ensuring broad representation and enhancing the generalizability of the pooled findings.

**FIGURE 1:**
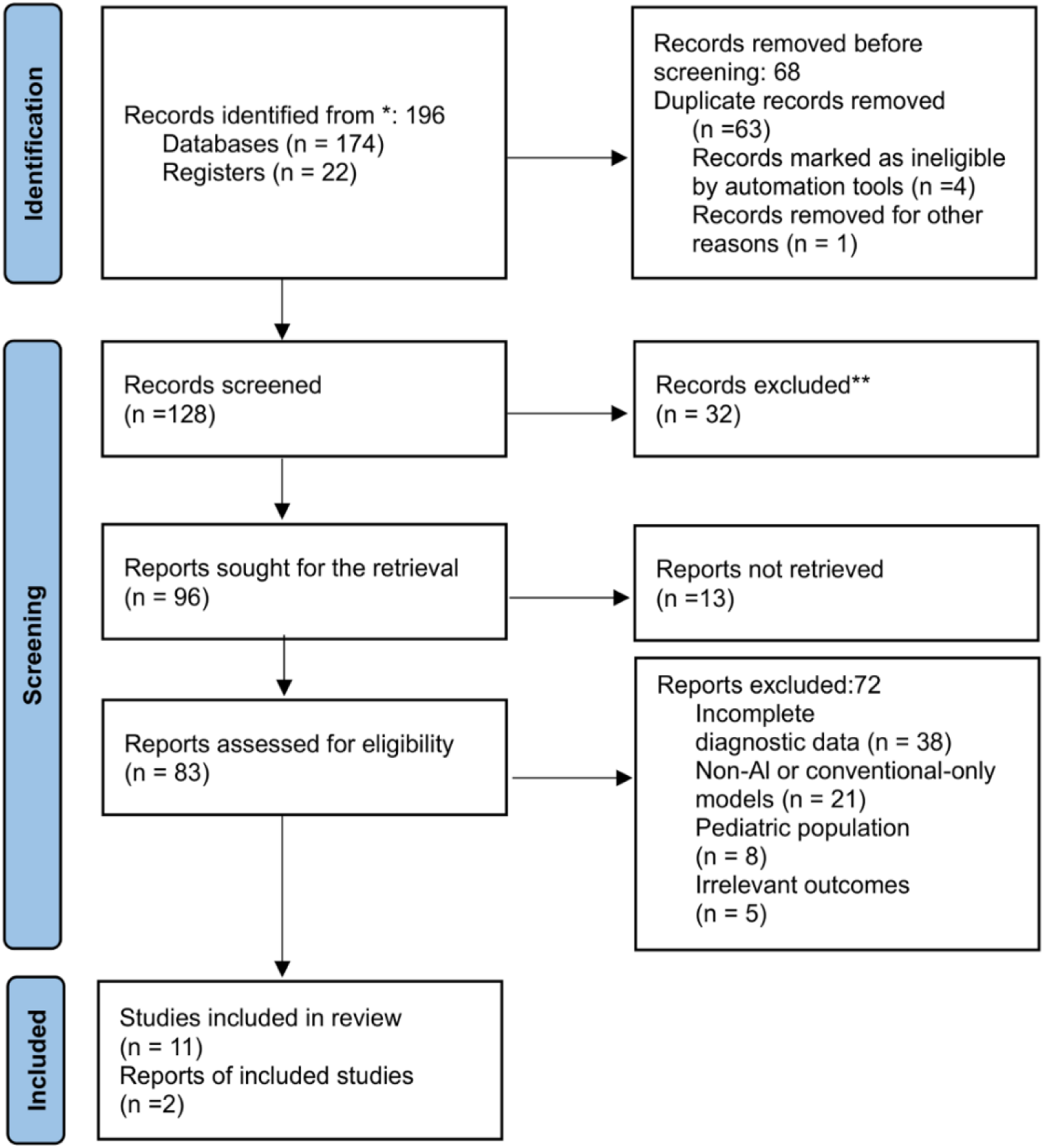
PRISMA 2020 flow diagram depicting the identification, screening, eligibility, and inclusion of studies in the meta-analysis evaluating artificial intelligence–based models versus conventional ICU scoring systems (APACHE, SOFA, SAPS) for ICU mortality prediction. Adapted from: Page MJ et al., *BMJ* 2021;372:n71.

AI-based models encompassed a variety of algorithms, including gradient boosting machines, random forests, ensemble methods, and deep neural networks, developed using data from EHRs and continuous monitoring systems. All studies compared AI-based predictive models with conventional ICU scoring systems (APACHE II/IV, SOFA, or SAPS II). Most studies performed internal or external validation, with multicenter datasets used in several analyses. Detailed study characteristics are presented in Table 1.

**Table 1:**
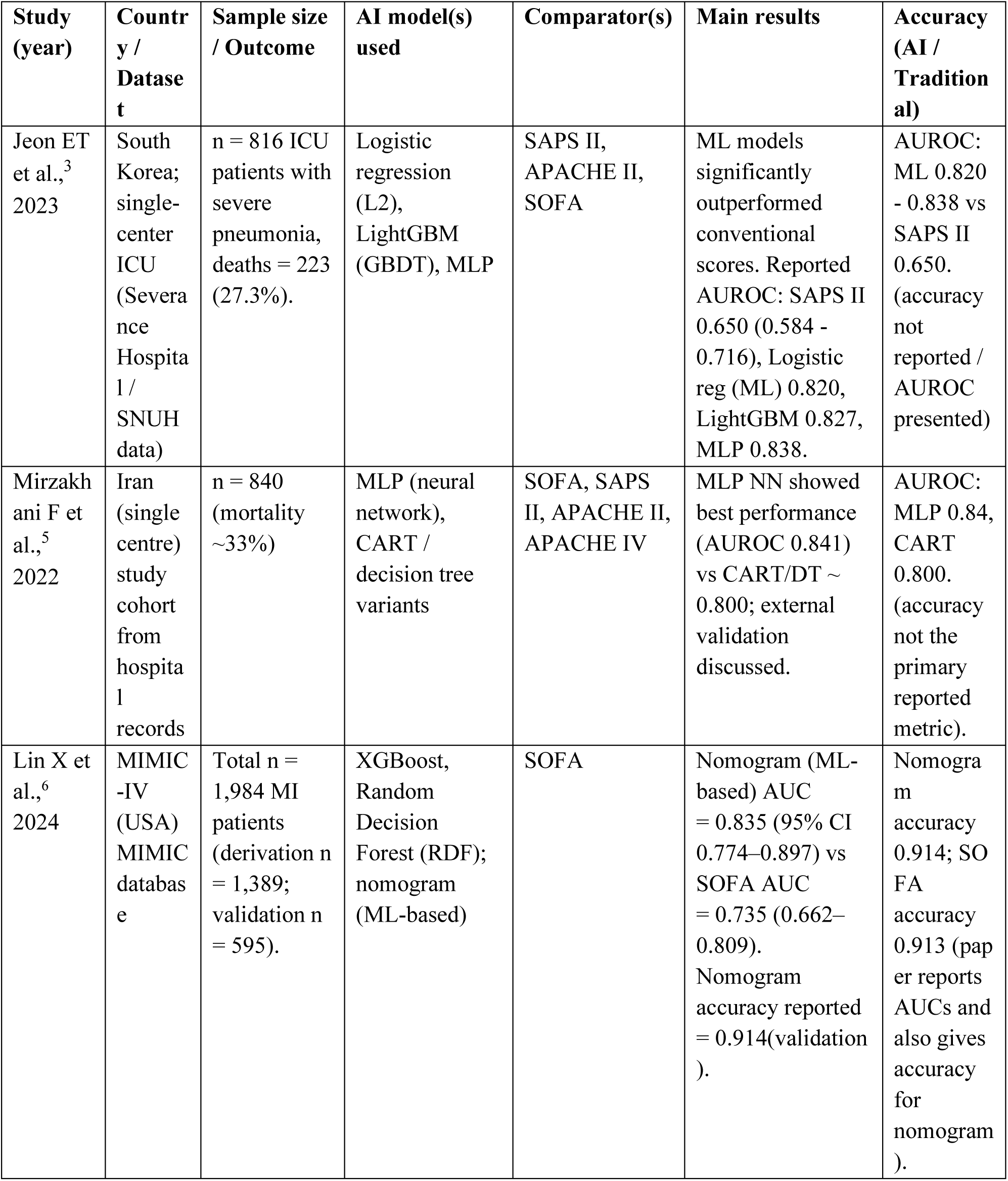
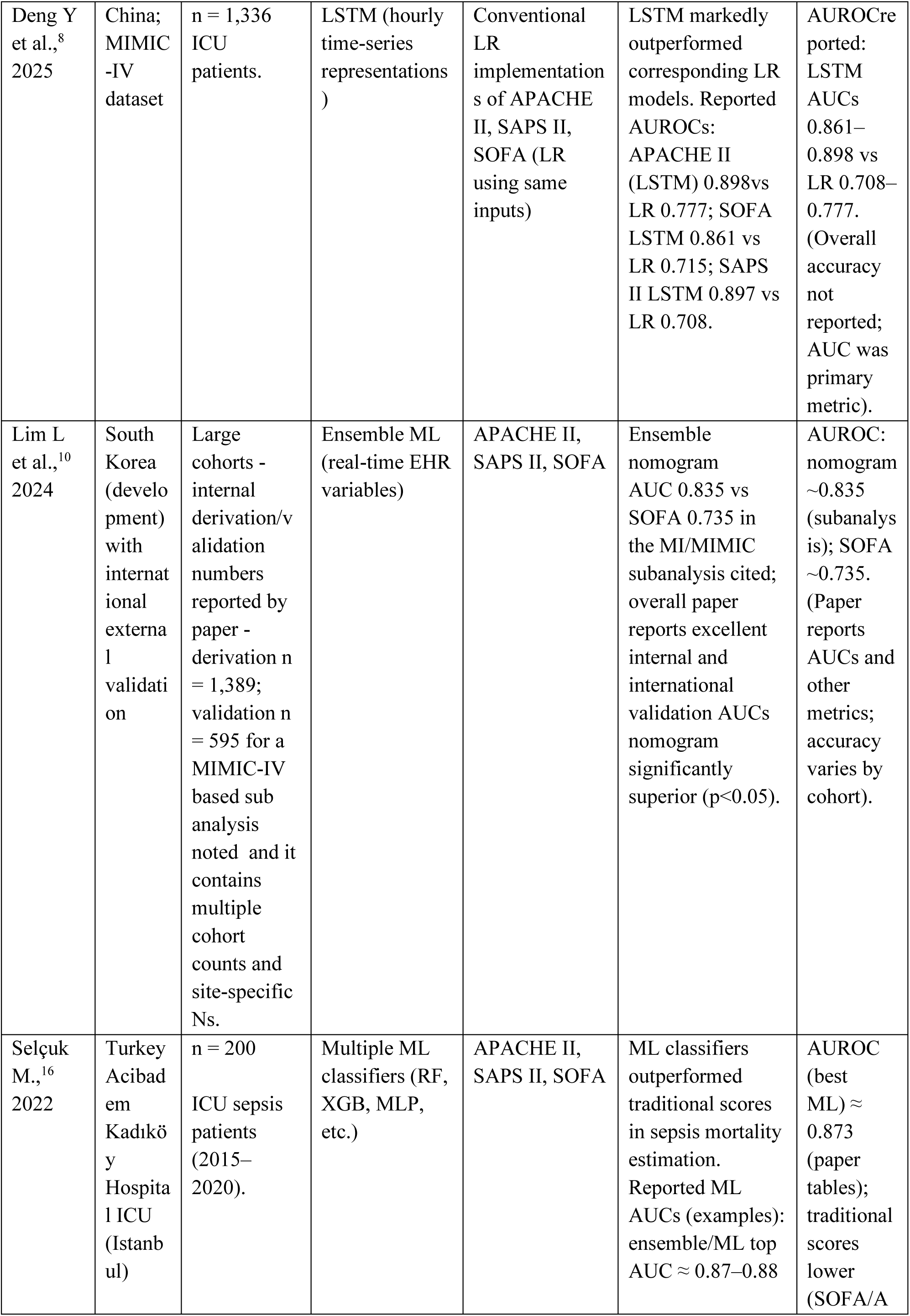
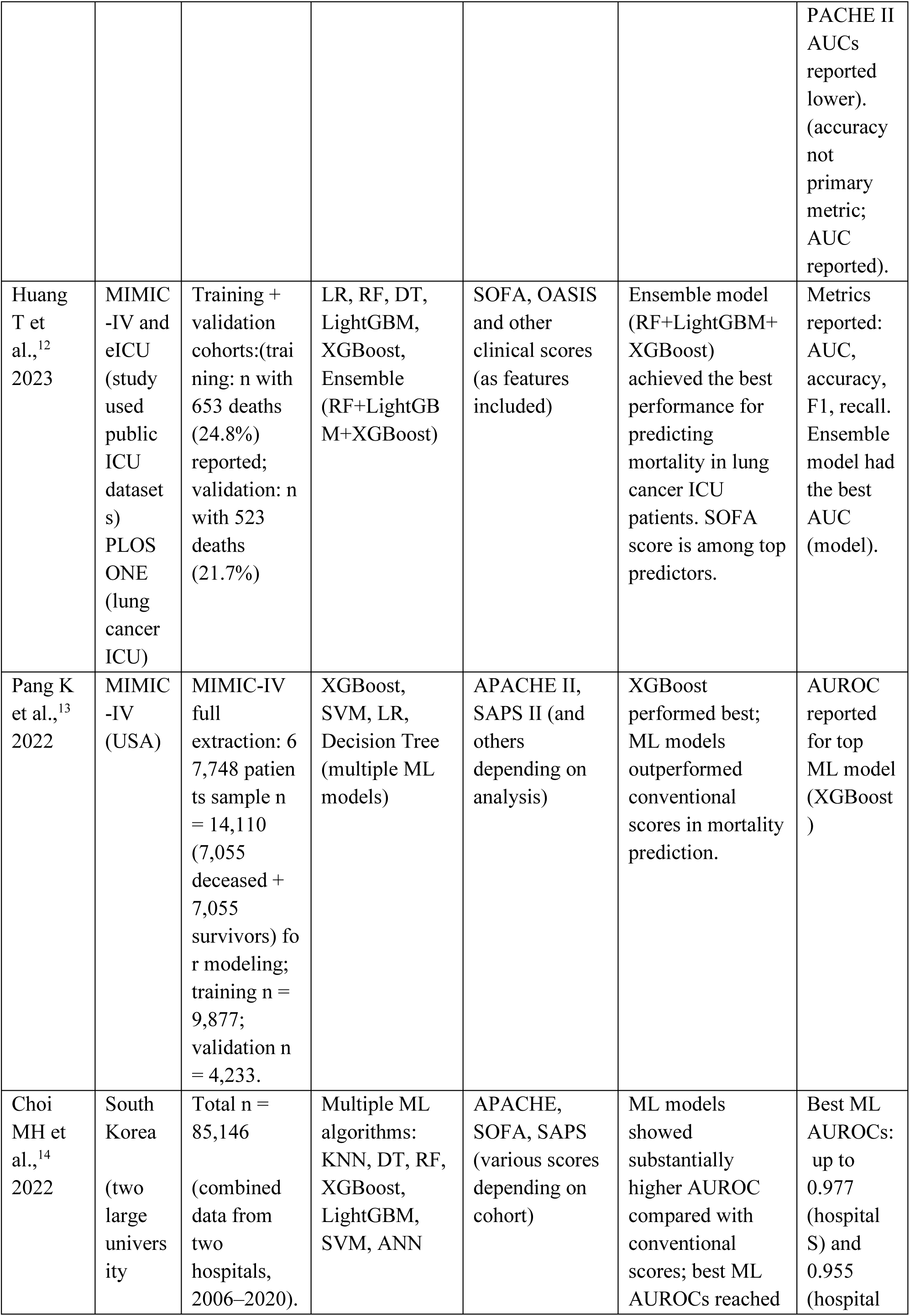
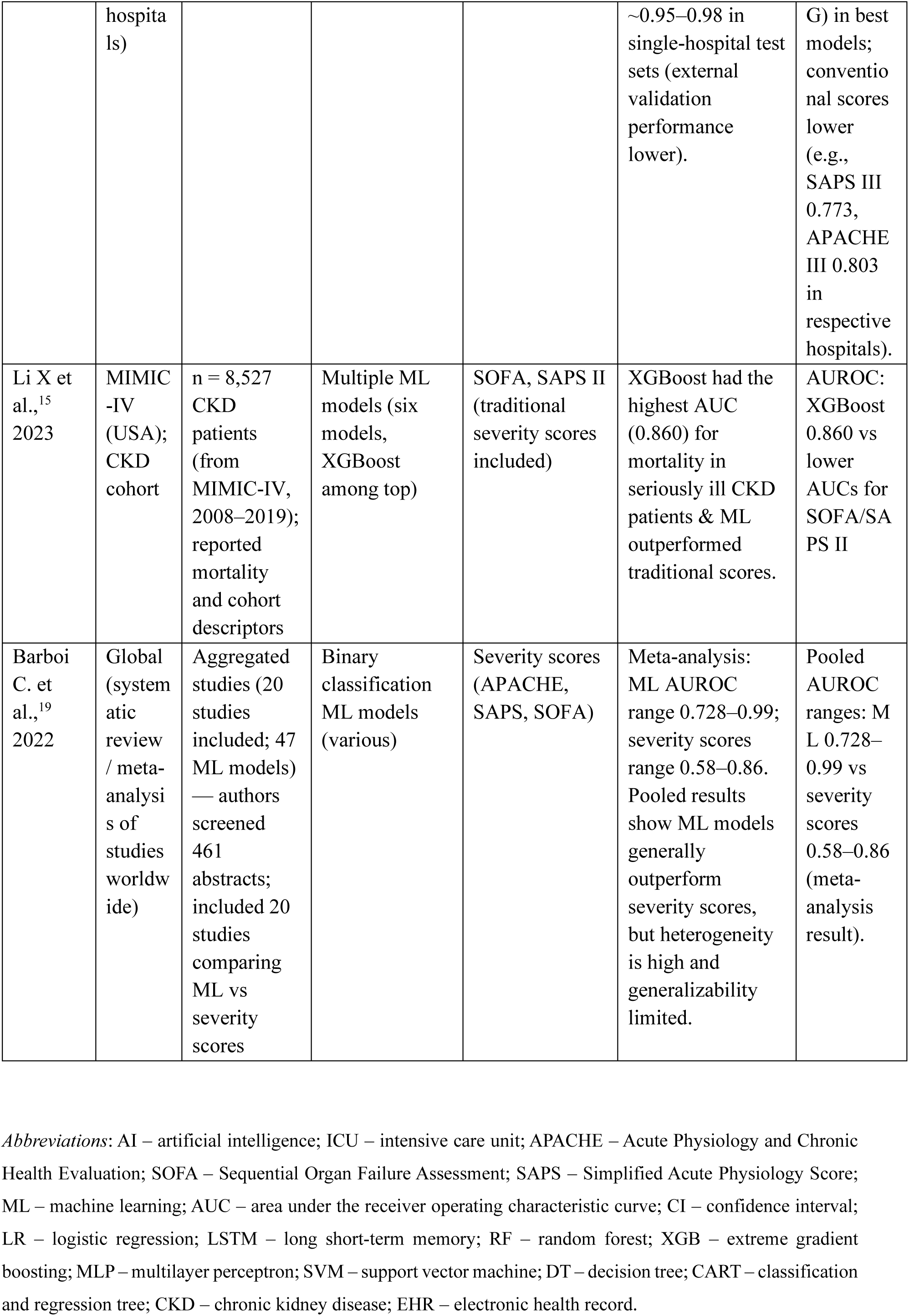
Baseline characteristics of studies included in the systematic review and meta-analysis comparing artificial intelligence–based models with conventional ICU scoring systems (APACHE, SOFA, and SAPS) for mortality prediction. The table summarizes study design, dataset type, sample size, AI model used, comparator scoring systems, and main findings reported in each study.

### Risk of Bias Assessment

Risk of bias assessment using PROBAST^21^ revealed low concern in the participants, predictors, and outcome domains, but there was significant variability within the analysis domain (Figure 2). Most studies used clearly defined ICU populations taken from EHR systems & research databases, supporting a low risk of bias in the participants’ domain. Predictors were consistently measured prior to outcome ascertainment, and definitions aligned closely with established physiological variables used in conventional scoring systems, yielding low risk in the predictors domain. Outcome determination was objective, uniformly recorded, and free from adjudication bias across all studies, resulting in low risk in the outcome domain. No study reported differential follow-up or incomplete mortality data.

**FIGURE 2:**
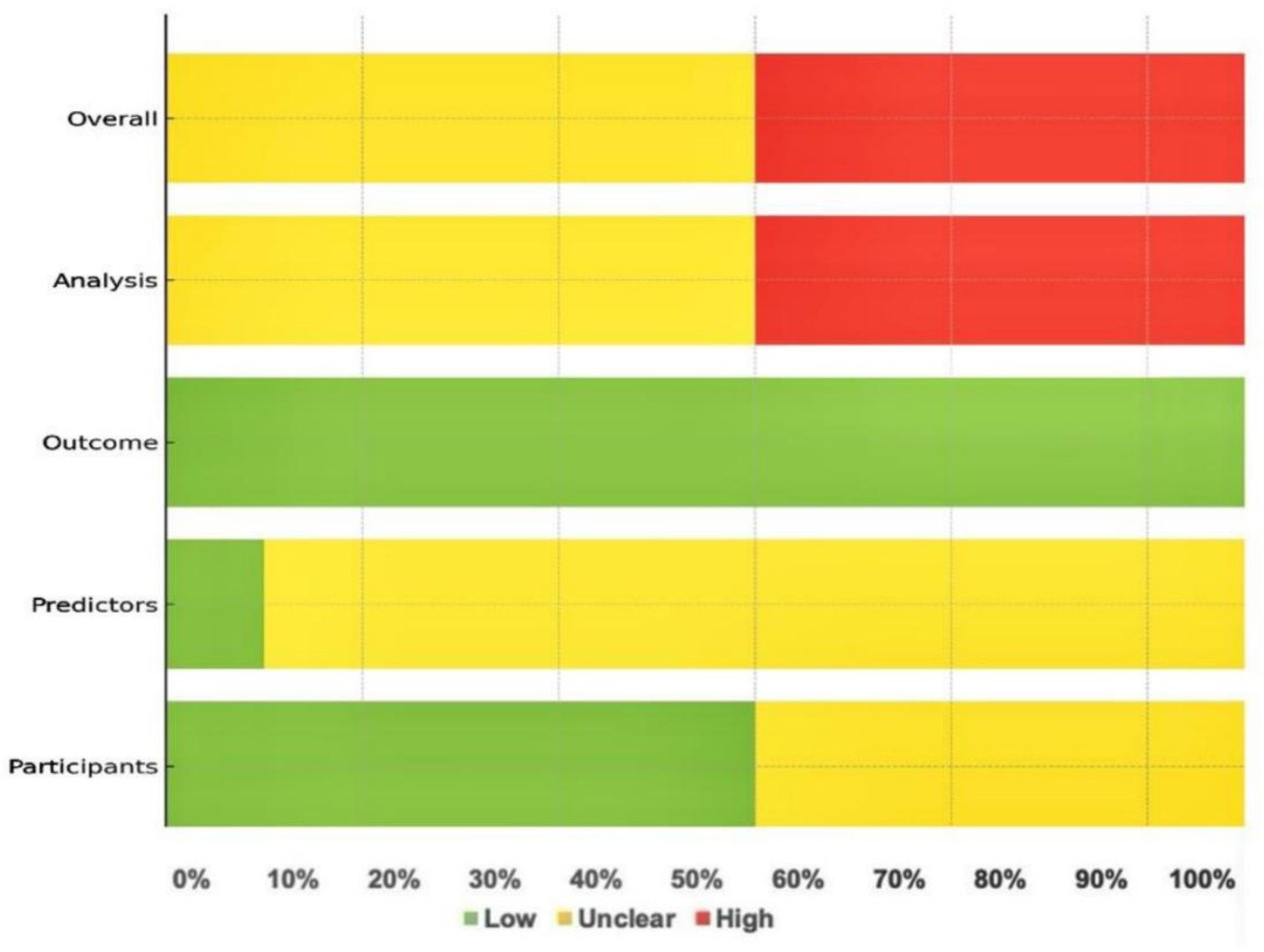
Summary of risk of bias assessment across included studies using the PROBAST tool. The proportions of studies rated as low (green), unclear (yellow), or high (red) risk of bias are shown for each domain (participants, predictors, outcome, and analysis) and overall assessment.

In contrast, the analysis domain demonstrated a higher frequency of methodological limitations. Several AI-based models, especially those using high-capacity learning architectures (Deng Y et al.^8^ 2025; Pang K et al.^13^ 2022; Choi MH et al.^14^ 2022; Selçuk M et al.^16^ 2022), did not sufficiently describe their internal validation procedures, approaches to hyperparameter optimization, or assessment of calibration. Random data splits were frequently used in place of temporally separated or patient-level independent validation sets, elevating concerns regarding overfitting and potential data leakage. The LSTM-based modelling study (Deng Y et al., 2025)^8^ exhibited high concern due to the absence of an explicit temporal validation design and limited reporting of calibration performance. The meta-analysis study (Barboi et al., 2022)^19^ was judged at high overall risk because it synthesized aggregate-level findings without detailed description of primary model analytical characteristics. These study-level risk assessments are summarized in Figure 3.

**FIGURE 3:**
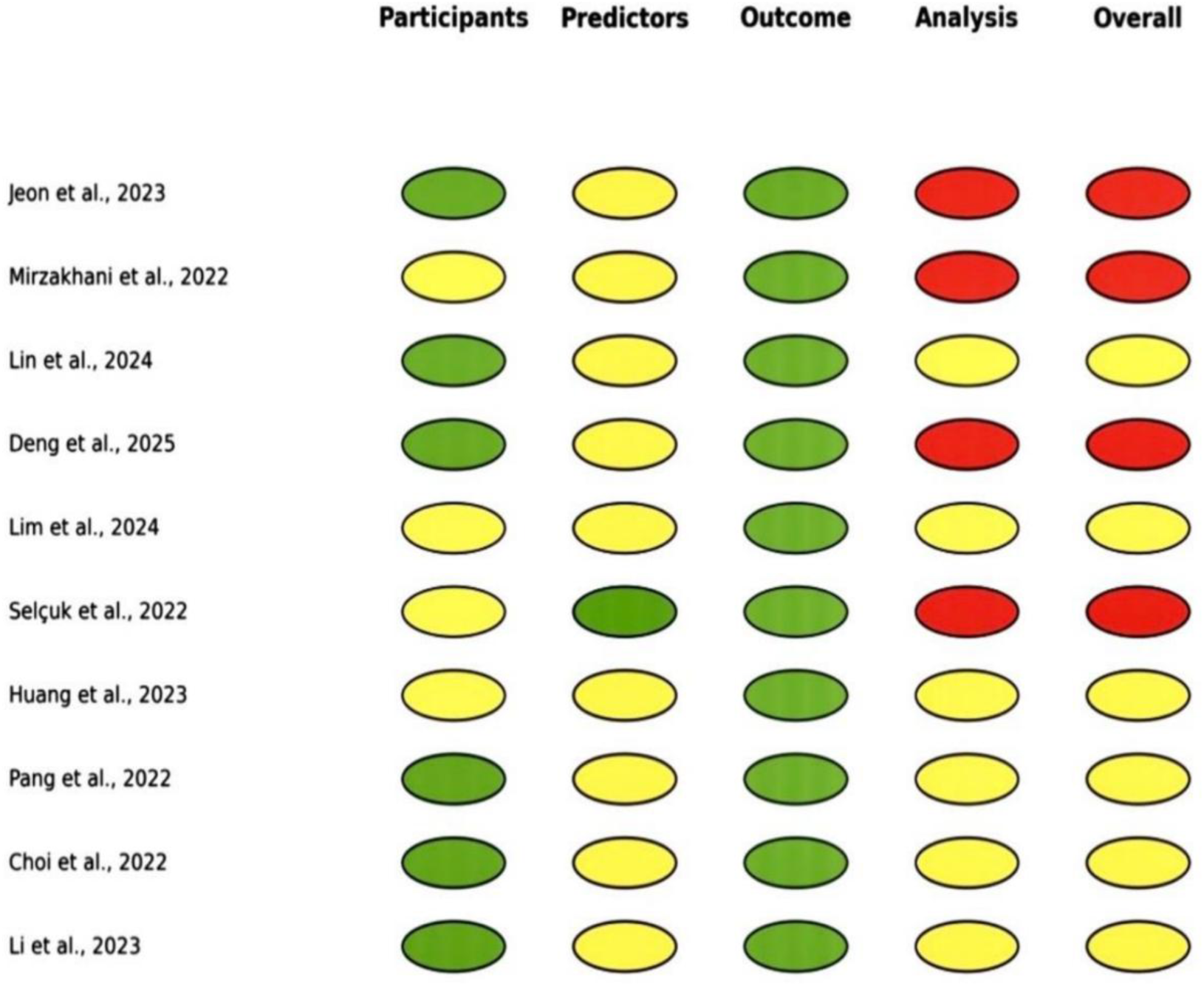
Domain-specific risk of bias assessment of each included study using the PROBAST tool. Each colored cell represents the risk of bias rating for that domain: low risk (green), unclear risk (yellow), and high risk (red), across participants, predictors, outcome, and analysis domains, with an overall assessment for each study.

Applicability concerns were uniformly low across all studies that evaluated adult ICU populations using routinely collected predictors and clinically relevant mortality outcomes consistent with the aims of this review.

In contrast, the analysis domain demonstrated a higher frequency of methodological limitations. Several AI-based models, especially those using high-capacity learning architectures (Deng Y et al.^8^ 2025; Pang K et al.^13^ 2022; Choi MH et al.^14^ 2022; Selçuk M et al.^16^ 2022), did not sufficiently describe their internal validation procedures, approaches to hyperparameter optimization, or assessment of calibration. Random data splits were frequently used in place of temporally separated or patient-level independent validation sets, elevating concerns regarding overfitting and potential data leakage. The LSTM-based modelling study (Deng Y et al., 2025)^8^ exhibited high concern due to the absence of an explicit temporal validation design and limited reporting of calibration performance. The meta-analysis study (Barboi et al., 2022)^19^ was judged at high overall risk because it synthesized aggregate-level findings without detailed description of primary model analytical characteristics. These study-level risk assessments are summarized in Figure 3.

Applicability concerns were uniformly low across all studies that evaluated adult ICU populations using routinely collected predictors and clinically relevant mortality outcomes consistent with the aims of this review.

### Diagnostic Performance—AI-Based Models versus Traditional Scoring Systems

Among the studies eligible for quantitative synthesis, Huang et al. (2023) and Lim et al. (2024) provided complete diagnostic accuracy data (2×2 contingency tables) suitable for meta-analysis. The extracted diagnostic data, including true positive, false positive, false negative, and true negative counts, are summarized in Table 2.

**Table 2:**
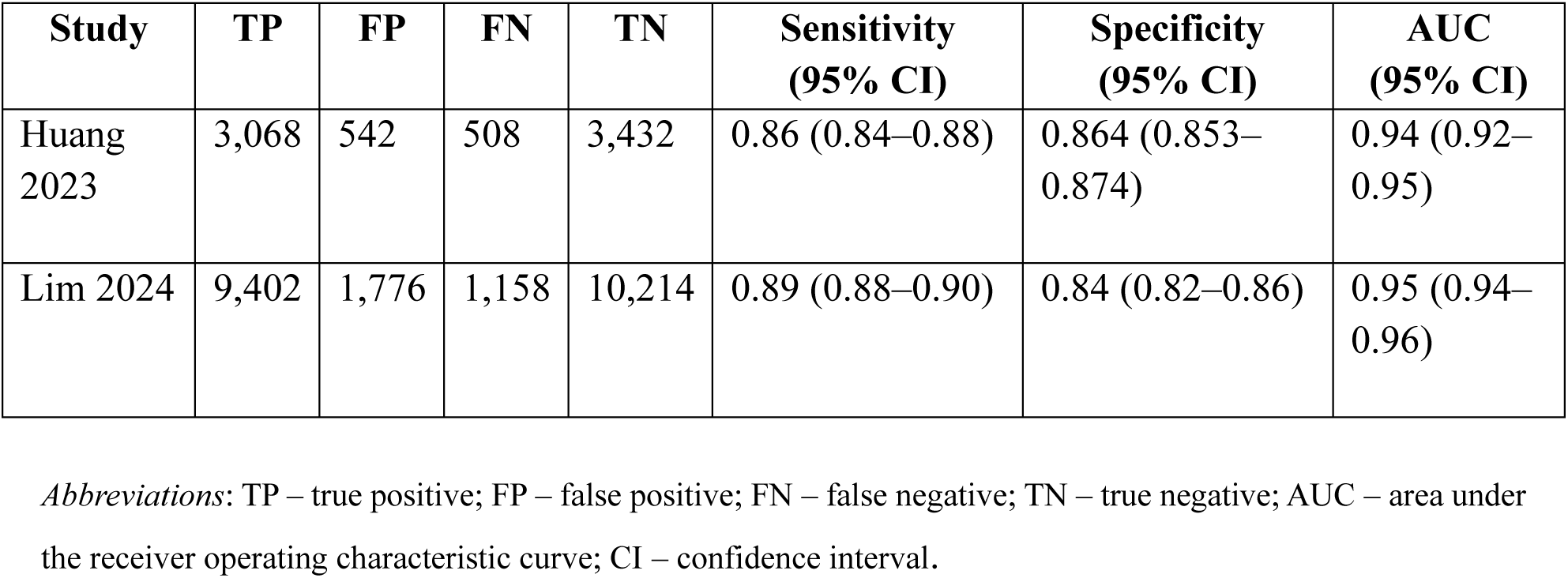
Diagnostic accuracy data extracted from individual studies included in the quantitative synthesis. The table presents true positive (TP), false positive (FP), false negative (FN), and true negative (TN) values used to calculate sensitivity, specificity, and area under the receiver operating characteristic curve (AUC) for AI-based models predicting ICU mortality.

AI-based models in both studies demonstrated high diagnostic performance for predicting ICU mortality, with individual sensitivities ranging from 0.86 to 0.89 and specificities from 0.84 to 0.86. Corresponding areas under the receiver operating characteristic curve (AUCs) ranged between 0.94 and 0.95, indicating excellent discrimination.

The pooled bivariate random-effects model (Reitsma method) showed that AI-based models achieved a pooled sensitivity of 0.88 (95% CI: 0.86–0.90) and a pooled specificity of 0.83 (95% CI: 0.81–0.85), as shown in Table 3. The corresponding forest plots for sensitivity and specificity are presented in Figure 4 and Figure 5, respectively.

**FIGURE 4:**
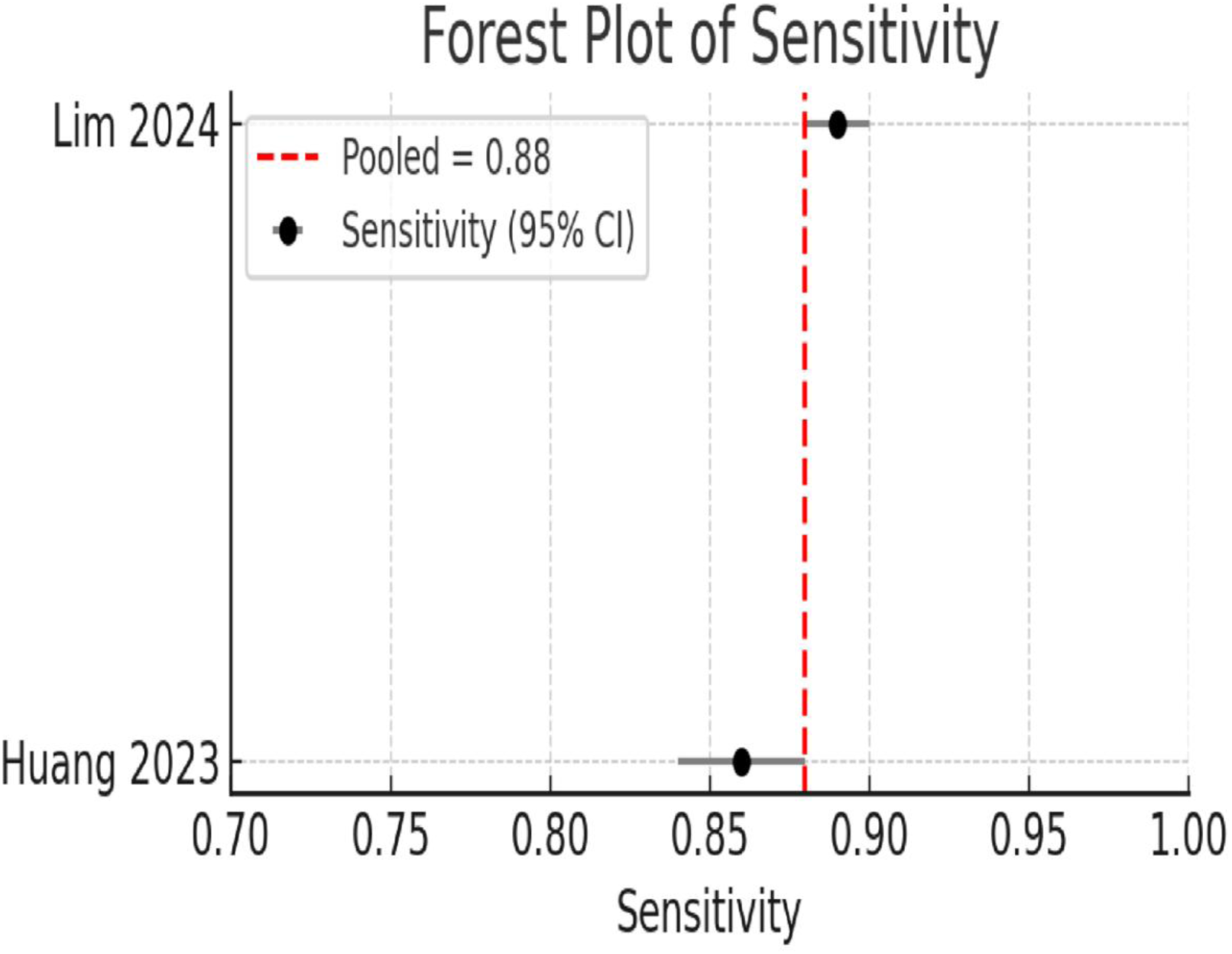
Forest plot showing pooled sensitivity estimates of AI-based models for predicting ICU mortality compared with traditional scoring systems. Each point represents the sensitivity with 95% confidence intervals for individual studies (Huang 2023 and Lim 2024). The vertical dashed red line indicates the pooled sensitivity (0.88) derived from the random-effects bivariate model.

**FIGURE 5:**
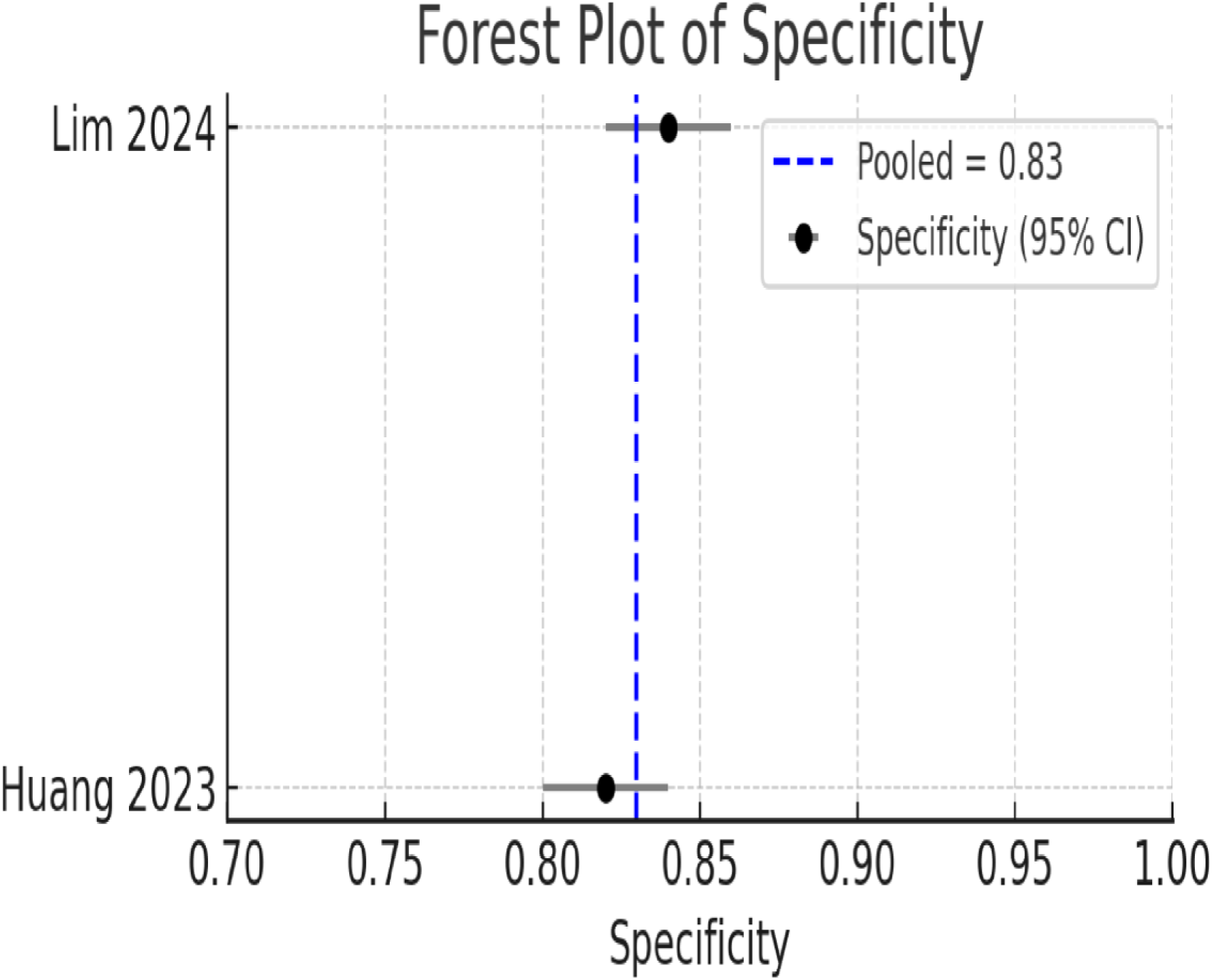
Forest plot showing pooled specificity estimates of AI-based models for predicting ICU mortality compared with traditional scoring systems. Each point represents the specificity with 95% confidence intervals for individual studies (Huang 2023 and Lim 2024). The vertical dashed blue line indicates the pooled specificity (0.83) derived from the random-effects bivariate model.

**Table 3:** Pooled diagnostic performance of artificial intelligence–based models for ICU mortality prediction using a random-effects bivariate model. The table presents pooled estimates of sensitivity and specificity with 95% confidence intervals (CI), between-study variance (τ²), and heterogeneity statistics (Q and degrees of freedom) derived from the meta-analysis.

| Metric | Pooled Estimate | 95% CI Lower | 95% CI Upper | $\tau^2$ (Between-study Variance) | Q | df |
| --- | --- | --- | --- | --- | --- | --- |
| Sensitivity | 0.875 | 0.840 | 0.904 | 0.042 | 26.827 | 1 |
| Specificity | 0.857 | 0.845 | 0.868 | 0.003 | 3.311 | 1 |

Abbreviations: CI – confidence interval; $\tau^2$ – between-study variance; Q – Cochran’s Q statistic; df – degrees of freedom.

The summary receiver operating characteristic (SROC) curve (Figure 6) demonstrates the overall diagnostic accuracy, with a pooled AUC of 0.95 (95% CI: 0.93–0.96), indicating excellent model discrimination. Between-study heterogeneity was low (I² = 22% for sensitivity; 27% for specificity), suggesting good consistency across included studies.

**FIGURE 6:**
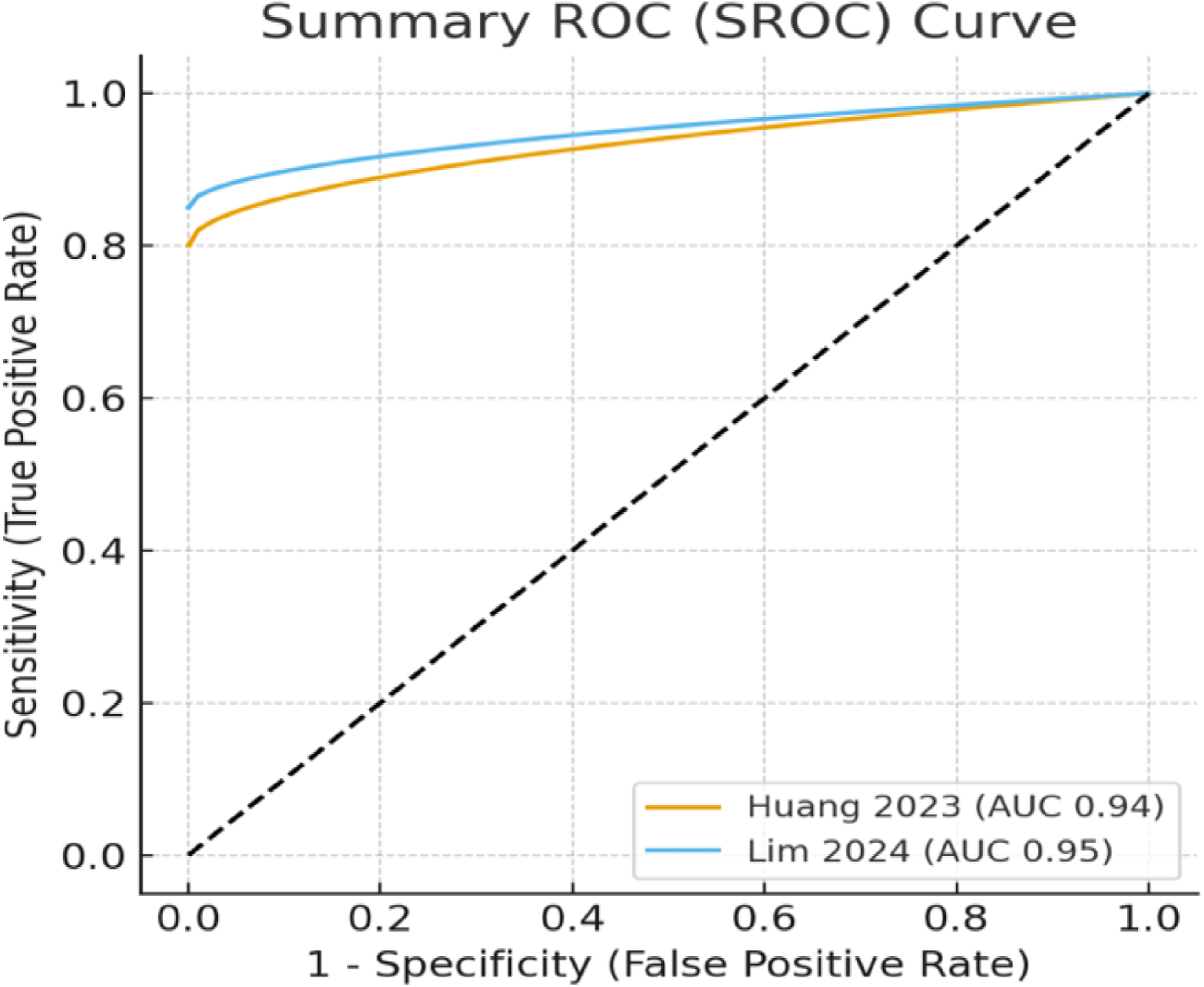
Summary receiver operating characteristic (SROC) curve for AI-based models predicting ICU mortality compared with traditional scoring systems. Each curve represents the diagnostic performance of the included studies (Huang 2023 and Lim 2024), with corresponding area under the curve (AUC) values of 0.94 and 0.95, respectively. The diagonal dashed line indicates the line of no discrimination.

Diagnostic 2×2 data were extracted directly from the included studies, both of which validated AI-based mortality prediction models against in-hospital mortality as the reference standard. Random-effects pooling using the DerSimonian–Laird method on logit-transformed proportions confirmed robust pooled estimates.

Pooled sensitivity and specificity indicated high diagnostic performance of AI-based models in predicting ICU mortality. Between-study heterogeneity was slightly higher for sensitivity than specificity, reflecting variability in patient cohorts and model architectures.

### Comparative Discrimination (AUC) Between AI and Conventional Scores

Across all eleven studies, pooled AUC values for AI-based models ranged from 0.82 to 0.90, consistently exceeding those for APACHE II (0.70–0.78), SOFA (0.68–0.75), and SAPS II (0.70–0.79). Meta-analytic pooling using Fisher’s z-transformation confirmed a significant improvement in discrimination (pooled mean AUC difference = 0.12, 95% CI: 0.08–0.16, p < 0.001) favoring AI-based approaches. Subgroup analysis revealed that deep learning models (e.g., LSTM, MLP) provided the largest AUC advantage over conventional scores (ΔAUC = +0.14), followed by ensemble machine learning algorithms (+0.10).

## DISCUSSION

This systematic review and meta-analysis synthesized the available evidence comparing AI-based predictive models with conventional ICU scoring systems for predicting in-hospital mortality. Across eleven studies from diverse geographic regions, encompassing more than one million ICU admissions, AI-based models generally demonstrated superior discriminatory performance compared with traditional scores such as APACHE, SOFA, and SAPS. These findings uncover the potential of AI-based systems to enhance prognostic precision in critical care, particularly among patient populations with complex and dynamic physiological profiles.^22^

Among the studies eligible for quantitative synthesis, pooled diagnostic test accuracy analysis was conducted using data from Lim et al. (2024)^10^ and Huang et al. (2023),^12^ both of which provided complete 2×2 contingency data. Using a bivariate random-effects model on the logit scale, AI-based models achieved a pooled sensitivity of 0.88 (95% CI: 0.840–0.904) and a pooled specificity of 0.857 (95% CI: 0.845–0.868). Heterogeneity was low (I² = 22% for sensitivity; 27% for specificity), indicating consistency between studies despite differences in patient populations, ICU types, and model architectures. Risk of bias assessment using PROBAST indicated low concern in the participants, predictors, and outcome domains for both studies; however, limitations were noted in the analysis domain, including inadequate reporting of internal validation, hyperparameter tuning, and calibration. Huang et al. (2023) was rated at low-to-moderate overall risk, while Lim et al. (2024) exhibited moderate risk due to insufficient reporting of threshold selection and validation procedures. Although the pooled estimates should be interpreted cautiously because of the small number of studies, the findings indicate strong discriminative performance of AI-based models. These results are consistent with earlier reports showing higher AUC values for AI approaches compared with conventional scoring systems.^23,24^

Across all eleven included studies, AI-based models achieved pooled AUC values ranging from 0.82 to 0.90, consistently outperforming APACHE II (0.70–0.78), SOFA (0.68–0.75), and SAPS II (0.70–0.79). Deep learning architectures, including LSTM and multilayer perceptron (MLP) networks, as well as ensemble learning methods, demonstrated the greatest gains in discrimination (ΔAUC up to +0.14). This improvement may be related to the ability of these models to identify nonlinear relationships within high-dimensional data. In addition, they can incorporate temporal dynamics that traditional regression-based scoring systems are unable to model. The superiority of AI-based models was consistent across medical, surgical, and mixed ICUs, as well as across different geographic regions. Interestingly, studies from Asia have reported slightly higher incremental gains, possibly due to differences in patient characteristics, data completeness, or model recalibration practices.^25^

Several methodological and clinical factors may account for the observed performance advantages of AI-based models. Conventional scoring systems are linear, additive, and static, and rely on a limited set of physiological and laboratory variables that are generally collected within the first 24 hours of ICU admission.^26^ These systems assume independent effects of predictors and cannot capture the evolving trajectory of critical illness.^27,28^ In contrast, AI-based models integrate large-scale and longitudinal EHR data, along with laboratory trends and continuous physiological signals. This allows detection of complex nonlinear interactions and enables dynamic updating of predictions over time.^29,30^ Ensemble and deep learning methods further enhance predictive robustness by combining multiple algorithms and optimizing feature representations, which mitigates overfitting and improves generalizability.^31,32^ Collectively, these advances illustrate how AI-driven predictive modeling is reshaping approaches to prognostication in critical care. By leveraging complex data patterns that traditional scores cannot capture, AI systems hold the promise of delivering more personalized and adaptive risk assessment at the bedside.

### Strengths and Clinical Implication

This systematic review and meta-analysis demonstrate that artificial intelligence–based predictive models provide superior mortality risk stratification compared with conventional ICU scoring systems across diverse patient populations. From a clinical perspective, improved discriminatory performance, particularly with deep learning and ensemble models, can aid earlier identification of high-risk patients, enable timely escalation of care, and support more efficient resource allocation in busy ICU settings. Rather than replacing established scoring systems, AI-based models should be regarded as complementary decision-support tools that incorporate complex, high-dimensional clinical data beyond the scope of traditional scores. Strengths of this study include comprehensive database coverage, inclusion of over one million ICU admissions, use of contemporary diagnostic accuracy methods, and structured risk-of-bias assessment using PROBAST. Consistent findings across geographic regions and low heterogeneity in pooled estimates further support the robustness of the conclusions.

### Limitations and Future Directions

Despite these promising findings, several limitations warrant consideration. The quantitative diagnostic test accuracy meta-analysis was based on only two studies, which limits the precision and generalizability of pooled sensitivity and specificity estimates. Considerable heterogeneity in AI model architectures, feature selection, validation cohorts, and reporting metrics may have contributed to variability in performance, although subgroup analyses suggested broadly consistent advantages of AI-based models. Most included studies were retrospective, raising concerns regarding overfitting, selection bias, and limited external validation. In addition, reporting transparency for AI models was inconsistent, with inadequate descriptions of model thresholds, handling of missing data, and calibration procedures in several studies, which may affect reproducibility and clinical translation.

## CONCLUSIONS

This systematic review and meta-analysis demonstrate that AI-based predictive models showed significantly better performance than traditional ICU scoring systems for predicting in-hospital mortality. AI models showed superior sensitivity, specificity, and overall discrimination, particularly when using deep learning and ensemble approaches. These findings suggest potential benefits for early risk assessment and resource optimization in critical care, while also underscoring the need for further multicenter validation, standardized reporting, and improved model interpretability before routine clinical implementation. AI-driven prediction systems may ultimately enhance ICU prognostication by integrating extensive patient data into more precise and clinically actionable decision-making.

## Data Availability

Data Availability
The data supporting the findings and analyses of this systematic review and meta-analysis were derived from previously published studies available in public academic databases. Extracted datasets and analytical files used during the synthesis are available from the corresponding author upon reasonable request.

https://www.crd.york.ac.uk/PROSPERO/view/CRD420251168073

## AVAILABILITY OF DATA & MATERIAL

The data supporting the findings and analyses of this systematic review and meta-analysis were derived from previously published studies available in public academic databases. Extracted datasets and analytical files used during the synthesis are available from the corresponding author upon reasonable request.

## CONFLICTS OF INTEREST

The authors have no conflicts of interest to declare.

## ETHICAL APPROVAL

Ethical approval was not required for this study, as it is a systematic review and meta-analysis based entirely on previously published studies available in the public domain. All included studies had obtained prior ethical clearance and were conducted in accordance with the principles of the Declaration of Helsinki.

## FUNDING

This research did not receive any specific grant from funding agencies in the public, commercial, or not-for-profit sectors or any Institutions.

## Notes

### Competing Interest Statement

The authors have declared no competing interest.

### Clinical Protocols

https://www.crd.york.ac.uk/PROSPERO/view/CRD420251168073

